# Changing dynamics of COVID-19 deaths during the SARS-CoV2 B.1.617.2 (Delta variant) outbreak in England and Wales: Reduced COVID-19 deaths among the Care Home residents

**DOI:** 10.1101/2022.02.09.22270705

**Authors:** Venkata R. Emani, Raghunath Reddy, Shaila R. Emani, Kartik K. Goswami, Kailash R. Maddula, Nikhila K. Reddy, Abirath S. Nakka, Nidhi K. Reddy, Dheeraj Nandanoor, Sanjeev Goswami

## Abstract

**Objectives:** Care Homes faced crisis during the SARS-CoV2 pandemic with increased mortality and various corrective interventions were undertaken. The impact of current SARS-CoV2 B.1.617.2 (Delta variant) outbreak on COVID-19 deaths in Care Homes is not completely known.

**Design:** Observational study

**Setting and Participants:** We included nationwide data of deaths from all causes and COVID-19 in England and Wales during March 7, 2020-November 26, 2021.

**Methods:** Impact of COVID-19 deaths in Care Homes, Hospital and Homes relative to total deaths from all causes was investigated.

**Results:** During March 7-August 28,2020; 15,414 (29.8%) of the 51,740 COVID-19 deaths occurred in Care Homes. During August 29, 2020-May 28, 2021, there were significant decrease in COVID-19 deaths in Care Homes ([19.2%]; RR 0.64; 95%CI 0.63-0.65;p<0.0001], which significantly declined further ([8.9%]; RR 0.46; 95%CI 0.43-0.48;p<0.0001) during May 29-November 26, 2021. During the March 7-August 28, 2020; 20.0% (15,514/76,906) of all Care Home deaths were caused by COVID-19, which significantly declined to 18.5% (RR 0.92; 95%CI 0.90-0.93; p<0.0001) during August 29, 2020-May 28, 2021 and declined further to 2.5% (RR 0.13; 95% CI 0.12-0.14;p<0.0001) during May 29-November 26, 2021. Increased percentages of COVID-19 deaths occurred in Hospitals ([72.3%]; RR 1.14; 95% CI 1.1-1.15;p<0.0001) and Homes ([6.3%]; RR 1.33; 95% CI 1.27-1.40; p<0.0001) during August 29, 2020-May 28, 2021 and further increases in the Hospitals ([80.8%]; RR 1.11; 95% CI 1.10-1.12; p<0.0001) and Homes ([8.8%]; RR 1.39; 95% CI 1.31-1.47; p<0.0001) during May 29-November 26, 2021.

**Conclusions and Implications:** There was a significantly lower number of COVID-19 deaths occurred in Care Homes and a significantly lower number of Care Homes deaths caused by COVID-19 during the Delta variant surge in England and Wales than the prior surges. Care Home residents are at highest risk for mortality, continuous monitoring and research on COVID-19 preventive interventions are essential.

## INTRODUCTION

During the COVID-19 global pandemic caused by SARS-CoV-2 (severe acute respiratory syndrome coronavirus 2), Care Homes and long-term care facilities (LTCFs) worldwide plunged into crisis which resulted in 25-50% of total COVID-19 deaths in various countries during the early part of the pandemic^1–6^. Numerous preventive measures were taken all over the world to reduce the COVID-19 infections among the Care Home residents and to improve the outcomes^7–18^.

The Office of National Statistics reported a sharp increase in the COVID-19 deaths among Care Home residents in England and Wales during the first two waves of SARS-CoV2 pandemic^19^. The COVID-19 surge since early June 2021, is predominantly due to the Delta variant (SARS-CoV2 B.1.617.2) and their outcomes of COVID-19 deaths among the Care Home residents is not known. We performed a comprehensive analysis of trends on the place of death of deaths from all causes and COVID-19 deaths in England and Wales during the Delta variant surge and compared them with prior surges since the beginning of SARS-CoV2 pandemic in March 2020.

### METHODS

In this observational study, we analyzed the nationwide data of the Care Home deaths in England and Wales in between March 7, 2020 to November 26, 2021 during the SARS-CoV2 pandemic.

An initial analysis was performed of the place of death of the total deaths from all causes in England and Wales since 2015 until November 26, 2021 using the data from UK Office of National Statistics^20,21^. The places of death (Home, Hospital, Care Home, Hospice, other community establishments and elsewhere) among these deaths were analyzed for every year from 2015 through November 26, 2021 to evaluate for yearly changes in the place of death particularly during the SARS-CoV2 pandemic since early 2020.

### Study period

The UK Office National Statistics started releasing weekly total deaths from all cases, weekly SARS-CoV2 (COVID-19) deaths and their place of death starting March 7, 2021 (week 11)^21^. We analyzed the data of the places of death of the 1,019,872 deaths from all causes and 153,179 COVID-19 deaths in England and Wales since March 7, 2020 (week 11) until November 26, 2021 (week 47). The comparative analysis of the place of death was performed for three time periods: 1) March 7-August 28, 2021(weeks 11-35) during the initial SARS-CoV2 surge, 2) August 29, 2020 through May 28, 2021 (weeks 2020-36 to 2021-21) from the start of the second surge until prior to the Delta variant surge and 3) May 29-November 26, 2021 during the Delta variant surge.

An initial analysis was performed of the impact of the COVID-19 deaths on the deaths from all causes in England and Wales during the three comparative periods during the study period.

### Analysis of the “place of death” of the weekly reported COVID-19 deaths during and prior to the Delta variant surge

A further analysis of the COVID-19 deaths occurring at the place of death (Home, Hospital, Care Home, Hospice, other community establishments and elsewhere) was performed among cumulative number of weekly COVID-19 deaths during the three comparative periods during the study (March 7-August 28, 2020; August 29,2020-May 28, 2021 and May 29-November 26, 2021). The percentage of total COVID-19 deaths occurring at various places of death were calculated for each study period and any changes in the place of deaths were compared with outcomes of each preceding comparative study period, and weekly changes were plotted on the graph ^21^.

### Analysis of the “place of death” of the weekly reported total deaths from all causes and the COVID-19 deaths during the study period

An analysis of the total deaths from all causes occurring at various places (Home, Hospital, Care Home, Hospice, other community establishments and elsewhere), total COVID deaths in England and Wales that occurred in various places (Home, Hospital, Care Home, Hospice, other community establishments and elsewhere) during the three comparative periods (March 7-August 28, 2020; August 29,2020-May 28, 2021 and May 29-November 26, 2021) were calculated. The percent of total deaths from all causes in England and Wales occurring at various places (Home, Hospital, Care Home, Hospice, other community establishments and elsewhere), the percentage of deaths occurring at various places (Home, Hospital, Care Home, Hospice ….) due to COVID-19, the percent of total deaths occurring at various places (Home, Hospital, Care Home, Hospice ….) contributing to total COVID-19 deaths from all causes was calculated and weekly changes of the outcomes were plotted on the graphs. The statistical comparisons of these outcomes were made in between each preceding study periods.

### Statistical analysis

The relative risk (RR), and 95% confidence interval and P value was calculated to compare the outcomes during the three study periods of the study from March 7, 2020 to November 26, 2021. RR is calculated using the formula, p value (<0.05) was considered significant for the differentiation between the groups.

## RESULTS

*Analysis of total deaths from all causes in* England and Wales *from 2015-2021*: Table 1 shows the yearly deaths in England and Wales from 2015 to 2020 and deaths during 2021 until November 26, 2021. The Table demonstrate the average percentage occurrence of yearly deaths with majority of deaths occurring in the Hospital setup (45.7%), followed by Homes (24.3%), Care Homes (22.0%), and Hospice (5.5%) during the years 2015-2019. There were 607,922 deaths recorded in England and Wales in 2020 which was an increase of 14.3% in comparison with average of deaths during 2015-2019. There were 528,420 total deaths reported during the year 2021 until November 26, 2021. In comparison with average deaths during the years 2015-2019, there were significantly increased percentages of deaths at Homes (27.4%; RR 1.16; p<0.0001) and Care Homes (27.4%; RR 1.016; p<0.0001) and significant decreased percentages of deaths occurred at Hospitals (42.4%; RR 0.90; p<0.0001) during the year 2020.

**Table 1:** Total yearly deaths from all causes in England and Wales and the place of deaths (2015-2021).

|  | Year |  |  |  |  |  |  |
| --- | --- | --- | --- | --- | --- | --- | --- |
| Place of death | 2015 | 2016 | 2017 | 2018 | 2019 | 2020 | 2021 |
| All deaths in England Wales (n= %) | 529,655 (100%) | 525048 (100%) | 533,253 (100%) | 541,589 (100%) | 530,841 (100%) | 607,922 (100%) | 528,420 (100%) |
| Hospital (acute or community, not psychiatric); (n= %) | 251,354 (47.5%) | 249794 (47.6%) | 249,256 (46.7%) | 24,9735 (46.1%) | 242,472 (45.7%) | 257,974 (42.4%) | 235,822 (44.6%) |
| Home (n= %) | 120,630 (22.8%) | 122714 (23.4%) | 125,623 (23.6%) | 128,388 (23.7%) | 128,918 (24.3%) | 166,576 (27.4%) | 151,986 (28.8%) |
| Care Home (n= %) | 117,213 (22.1) | 112023 (21.3%) | 116,724 (21.9%) | 119,629 (22.1%) | 116,698 (22.0%) | 141,406 (23.3%) | 103,692 (19.6%) |
| Hospice (n= %) | 28,684 (5.4%) | 28635 (5.5%) | 29,761 (5.6%) | 30,396 (5.6%) | 29,456 (5.5%) | 26,207 (4.3%) | 22,189 (4.2%) |
| Elsewhere (n= %) | 10,011 (1.9%) | 10125 (1.9%) | 10,154 (1.9%) | 11,637 (2.1%) | 11,442 (2.2%) | 13,564 (2.2%) | 12,858 (2.4%) |
| Other communal establishment (n= %) | 1,763 (0.3%) | 1757 (0.3%) | 1,735 (0.3%) | 1,804 (0.3%) | 1,855 (0.3%) | 2,195 (0.4%) | 1,873 (0.4%) |
| **Years 2015-2019 (average) and 2020 statistical comparisons** |  |  |  |  |  |  |  |
| Place of death | 2015-2019 (average) | 2020 | RR; 95% CI; p value |  |  |  |  |
| All deaths in England Wales (n= %) | 532,077 (100%) | 607,922 (100%) |  |  |  |  |  |
| Hospital (acute or community, not psychiatric); (n= %) | 248,522 (46.7%) | 257,974 (42.4%) | RR 0.9085; 95% CI 0.9048 to 0.9123; P < 0.0001 |  |  |  |  |
| Home (n= %) | 125,255 (23.5%) | 166,576 (27.4%) | RR 1.1640; 95% CI 1.1566 to 1.1714; P < 0.0001 |  |  |  |  |
| Care Home (n= %) | 116,457 (21.9%) | 141,406 (23.3%) | RR 1.0627; 95% CI 1.0555 to 1.0700; P < 0.0001 |  |  |  |  |
| Hospice (n= %) | 29,386 (5.5%) | 26,207 (4.3%) | RR 0.7806; 95% CI 0.7680 to 0.7933; P < 0.0001 |  |  |  |  |
| Elsewhere (n= %) | 10,674 (2.0%) | 13,564 (2.2%) | RR 1.1122; 95% CI 1.0847 to 1.1405; P < 0.0001 |  |  |  |  |
| Other communal establishment (n= %) | 1,783 (0.3%) | 2,195 (0.4%) | RR 1.0775; 95% CI 1.0123 to 1.1468; P = 0.0190 |  |  |  |  |

During the study period (March 7, 2020-November 26, 2021) there were a total of 1,025,282 deaths from all causes and 153,179 deaths reported due to COVID-19 in England and Wales.

*COVID-19 deaths and the impact on deaths from all causes in* England and Wales (Table 4): During the March 7-August 28, 2020 period, there were 299,844 deaths from all causes and 51,740 (17.3%) deaths due to COVID-19, and during the August 29,2020-May 28, 2021 period there was a significant increase in the percentage of COVID-19 deaths to 19.0% of deaths from all causes (86,627 COVID-19 deaths/456,847 deaths from all causes; RR 1.09; p<0.0001). During the Delta variant surge (May 29-November 26,2021) the percentage of COVID-19 deaths decreased to 5.5% of deaths from all causes (14,812 COVID-19 deaths/268,591 deaths from all causes; RR 0.29; p<0.0001) in comparison with the prior period.

### Demographic changes in the place of COVID-19 deaths

Table 2 demonstrates the distribution of the 153,179 total COVID-19 deaths in England and Wales during the study period and percentage of the COVID-19 deaths occurring at Homes, Hospitals, Care Homes, Hospice, other community establishments and elsewhere. There was a total of 51,740 COVID-19 deaths during March 7-August 29, 2020; 86,627 COVID-19 deaths during August 29-May 28, 2021 and 14,812 COVID-19 deaths during the May 29-November 26, 2021 study period.

**Table 2:** Total SARS-CoV2 deaths in England and Wales and the place of deaths during the study period (March 7, 2020 -November 26, 2021)

|  | Study period |  |  |  |  |
| --- | --- | --- | --- | --- | --- |
| **Place of Death**<br>(% of COVID-19 deaths occurring at each place) | **March 7-Aug 28, 2020 (n= %)** | **August 29, 2020-May 28, 2021 (n= %)** | **RR; 95% CI, p value\*** | **May 29-November 26, 2021 (n= %)** | **RR; 95% CI, p value\*** |
| **COVID-19 deaths (n=153,179)** | 51,740 (100%) | 86,627 (100%) |  | 14,812 (100%) |  |
| Hospital (acute or community, not psychiatric) | 32,731 (63.3%) | 62,653 (72.3%) | RR 1.1433; 95% CI 1.1345 to 1.1522; P < 0.0001 | 11,968 (80.8%) | RR 1.1172; 95% CI 1.1073 to 1.1271; P < 0.0001 |
| Care Home | 15,414 (29.8%) | 16,603 (19.2%) | RR 0.6433; 95% CI 0.6312 to 0.6557; P < 0.0001 | 1,312 (8.9%) | RR 0.4622; 95% CI 0.4381 to 0.4875; P < 0.0001 |
| Home | 2,432 (4.7%) | 5,451 (6.3%) | RR 1.3387; 95% CI 1.2778 to 1.4025; P < 0.0001 | 1,297 (8.8%) | RR 1.3916; 95% CI 1.3132 to 1.4746; P < 0.0001 |
| Hospice | 730 (1.4%) | 1,315 (1.5%) | RR 1.0759; 95% CI 0.9835 to 1.1770; P = 0.1103 | 129 (0.9%) | RR 0.5737; 95% CI 0.4792 to 0.6869; P < 0.0001 |
| Other communal establishment | 228 (0.4%) | 292 (0.3%) | RR 0.7649; 95% CI 0.6435 to 0.9093; P = 0.0024 | 24 (0.2%) | RR 0.4807; 95% CI 0.3172 to 0.7286; P = 0.0006 |
| Elsewhere | 205 (0.4%) | 313 (0.4%) | RR 0.9119; 95% CI 0.7649 to 1.0872; P = 0.3040 | 82 (0.6%) | RR 1.5322; 95% CI 1.2022 to 1.9527; P = 0.0006 |

During the March 7-August 28, 2020 period (first wave), of the total 51,740 COVID-19 deaths, 32,731 (63.3%) occurred in the Hospitals; 15,414 (29.8%) COVID-19 deaths in Care Homes; 2,442 (4.7%) COVID-19 deaths at Homes, and 730 (1.4%) COVID-19 deaths in Hospice. There was a significant decrease in the percentage of COVID-19 deaths occurring in Care Homes of the total COVID-19 deaths [16,603 (19.2%); RR 0.64; p<0.0001] during August 29, 2020-May 28, 2021 period. The COVID-19 deaths occurring in Care Homes decreased significantly (1,312 [8.9%]; RR 0.46; p<0.0001) during the Delta variant surge (May 29-November 26, 2021) compared to prior period. The COVID-19 deaths occurring in the Hospitals increased to 72.3% of total COVID-19 deaths (n=62,653; RR 1.14; p < 0.0001) during the August 29,2020-May 28, 2021 and further significantly increased to 80.8% of total COVID-19 deaths (n=11,968; RR 1.11; p<0.0001) during the Delta variant surge (May 29-Novemver 26, 2021). Similarly, COVID-19 deaths occurring at Homes showed a significant increase (5,451 [6.3%]; RR 1.33; p<0.0001) during August 29, 2020-May 28, 2021 and further significantly increased (1,297 [8.8%]; RR 1.39; p<0.0001) during the Delta variant surge (May 29-November 26, 2021). Figure 1 illustrates that, up to 43.6% of weekly COVID-19 deaths occurred in the Care Homes during the initial surge (March-August 2020), during the second surge after January 2021 period the percent of COVID-19 deaths occurring in Care Homes decreased to 23.6% of weekly total COVID-19 deaths. During the Delta variant surge, the percent of the weekly COVID-19 deaths occurring in the Care Homes decreased to a maximum of 10.8% total COVID-19 deaths. Figure 4 also illustrate the percentage of weekly COVID-19 deaths occurring in the hospital declined to lowest level of 50.1% of total COVID-19 deaths during the first wave (coinciding with increased COVID-19 deaths occurring in Care Homes) and during the second surge there was a steady increase in the percentage of COVID-19 deaths occurring in the Hospitals and during the Delta variant surge, this was increased to as high as 85.3% of total weekly COVID-19 deaths occurring in the hospitals.

**Figure 1:**
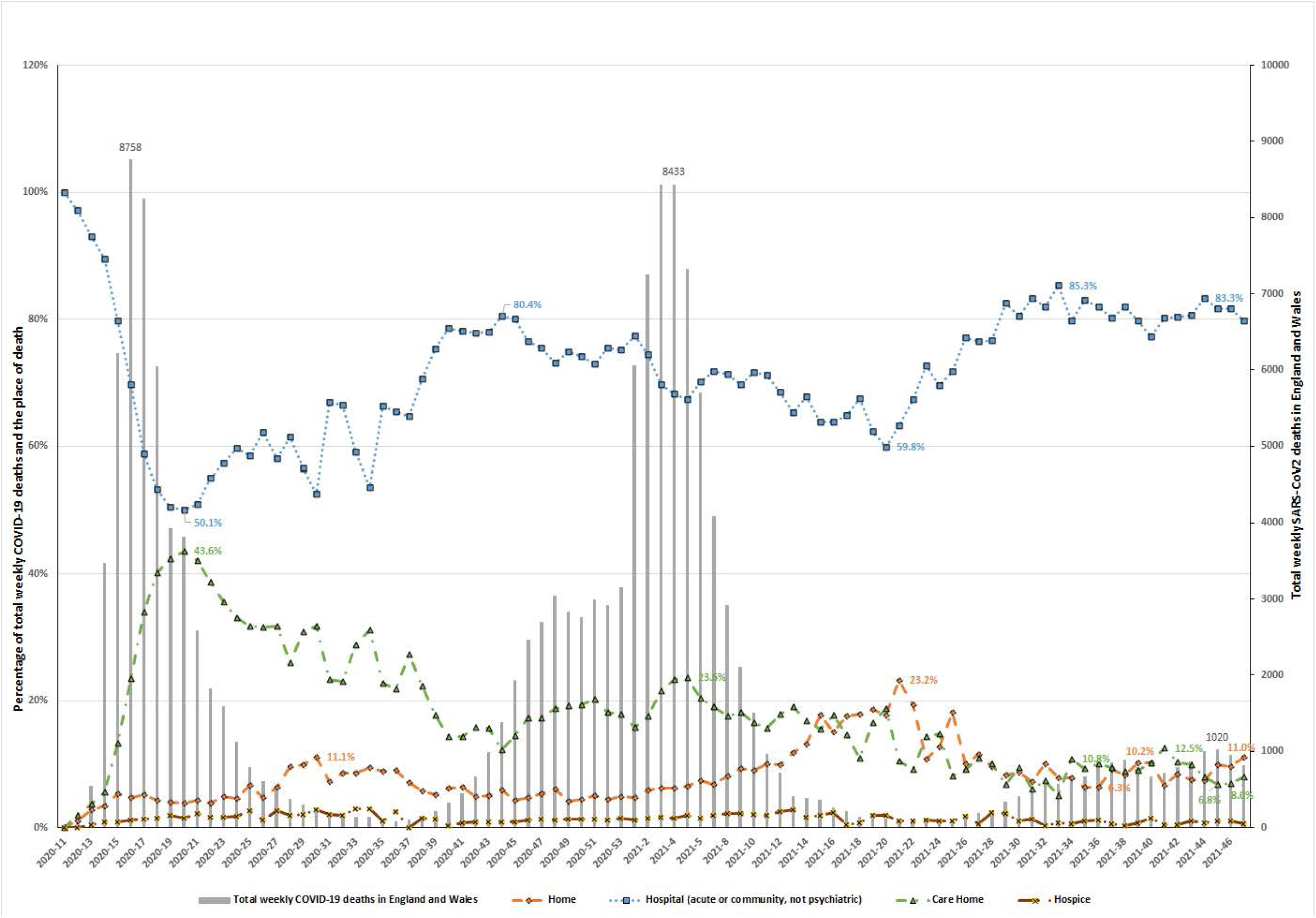
Shows the proportion of weekly COVID-19 deaths based on place of death (primary axis) and total weekly deaths (secondary axis). Since the Delta variant surge in June 2021, the proportion of COVID-19 deaths occurring in Hospitals increased significantly. There is also significant decline in the proportion of weekly COVID-19 deaths in Care Homes during the Delta variant surge compared to the prior surges. During the 1^st^ surge, up to 43.6% of weekly COVID-19 deaths occurred at Care Homes, with associated decreases in deaths occurring in the Hospital setup.

### Deaths from all causes at Care Homes; the percentage deaths from all causes occurring at the Care Homes

Table 1 shows the percentage of total deaths each year occurring in Care Homes, on comparison with 2019, during the year 2020 there were a significant increased percentage of deaths in Care Homes (141,406 [23.3%] in 2020 vs 116,698 [22.0%] in 2019; RR 1.05; 95% CI 1.05 to 1.06; P<0.0001). Table 3 shows the Care Homes deaths from all causes (n=220,092) during the study period (March 7, 2020-November 26, 2021). During March 7-August 29, 2020 (first wave), of the total 299,844 deaths from all causes; 76,906 (25.6%) occurred in Care Homes. During the August 29, 2020-May 28, 2021 there was a significant decrease in Care Home deaths (n=89,954; [19.7%]; RR 0.76; p<0.0001) of the total deaths from all causes (n=456,847). During the May 29-November 26, 2021 study period coinciding with the Delta variant surge, the percentage of Care Home deaths remain the same (53,232 deaths of 268,591 total deaths [9.8%]; RR 1.00; P=0.18). Figure 2 demonstrate the higher percentage of weekly deaths up to 36.0% of total all cause deaths occurring during the initial surge in Care Homes, observed coinciding with peak cases during the surge.

**Table 3:** Total deaths (all causes) at the places of deaths; and the percentage of deaths due to COVID-19 at the places of deaths in England and Wales during the study period (March 7, 2020 -November 26, 2021).

|  | Study period |  |  |  |  |
| --- | --- | --- | --- | --- | --- |
| Characteristics of deaths | March 7-August 28, 2020 | August 29, 2020-May 28, 2021 | RR; 95% CI, p value* | May 29-November 26, 2021 | RR; 95% CI, p value* |
| **Total deaths from all causes (n=1,025,282)** | **299,844 (100%)** | **456,847 (100%)** |  | **268,591 (100%)** |  |
| **Percentage of deaths (all causes) and place of deaths (n= %)** |  |  |  |  |  |
| Hospital deaths (n=442,875) | 120,273 (40.1%) | 206,411 (45.2%) | RR 1.1264; 95% CI 1.1203 to 1.1325; P < 0.0001 | 116,191 (43.3%) | RR 0.9575; 95% CI 0.9523 to 0.9626; P < 0.0001 |
| Home deaths (n=291,705) | 82,713 (27.6%) | 129,926 (28.4%) | RR 1.0310; 95% CI 1.0234 to 1.0386; P < 0.0001 | 79,066 (29.4%) | RR 1.0351; 95% CI 1.0274 to 1.0428; P < 0.0001 |
| Care Home deaths (n=220,092) | 76,906 (25.6%) | 89,954 (19.7%) | RR 0.7677; 95% CI 0.7612 to 0.7742; P < 0.0001 | 53,232 (19.8%) | RR 1.0065; 95% CI 0.9969 to 1.0163; P = 0.1831 |
| Hospice deaths (n=42,704) | 12,261 (4.1%) | 18,317 (4.0%) | RR 0.9805; 95% CI 0.9588 to 1.0027; P = 0.0851 | 12,126 (4.5%) | RR 1.1260; 95% CI 1.1010 to 1.1516; P < 0.0001 |
| Elsewhere (n=24,088) | 6,441 (2.1%) | 10,612 (2.3%) | RR 1.0814; 95% CI 1.0488 to 1.1150; P < 0.0001 | 7,035 (2.6%) | RR 1.1276; 95% CI 1.0945 to 1.1616; P < 0.0001 |
| Other communal establishment (n=3,818) | 1,250 (0.4%) | 1,627 (0.4%) | RR 0.8543; 95% CI 0.7937 to 0.9195; P < 0.0001 | 941 (0.4%) | RR 0.9837; 95% CI 0.9080 to 1.0658; P = 0.6885 |
| **Percentage of deaths caused by COVID-19 (deaths due to COVID-19 at place of death/deaths from all causes at place of death); (n= %)** |  |  |  |  |  |
| % of Hospital deaths due to COVID-19 (n=107,352) | 32,731 (27.2%) | 62,653 (30.4%) | RR 1.1154; 95% CI 1.1028 to 1.1281; P < 0.0001 | 11,968 (10.3%) | RR 0.3393; 95% CI 0.3332 to 0.3456; P < 0.0001 |
| % of Home deaths due to COVID-19 (n=9,180) | 2,432 (2.9%) | 5,451 (4.2%) | RR 1.4269; 95% CI 1.3614 to 1.4955; P < 0.0001 | 1,297 (1.6%) | RR 0.3910; 95% CI 0.3683 to 0.4151; P < 0.0001 |
| % of Care Home deaths due to COVID-19 (n=33,329) | 15,414 (20.0%) | 16,603 (18.5%) | RR 0.9209; 95% CI 0.9029 to 0.9392; P < 0.0001 | 1,312 (2.5%) | RR 0.1335; 95% CI 0.1264 to 0.1411; P < 0.0001 |
| % of Hospice deaths due to COVID-19 (n=2,174) | 730 (6.0%) | 1,315 (7.2%) | RR 1.2058; 95% CI 1.1047 to 1.3161; P < 0.0001 | 129 (1.1%) | RR 0.1482; 95% CI 0.1239 to 0.1773; P < 0.0001 |
| % of deaths Elsewhere-due to COVID-19 (n=600) | 205 (3.2%) | 313 (2.9%) | RR 0.9267; 95% CI 0.7792 to 1.1021; P = 0.3895 | 82 (1.2%) | RR 0.3952; 95% CI 0.3105 to 0.5030; P < 0.0001 |
| % of deaths-Other communal establishment-due to COVID-19 (n=544) | 228 (18.2%) | 292 (17.9%) | RR 0.9839; 95% CI 0.8412 to 1.1509; P = 0.8396 | 24 (2.6%) | RR 0.1421; 95% CI 0.0945 to 0.2138; P < 0.0001 |

**Figure 2:**
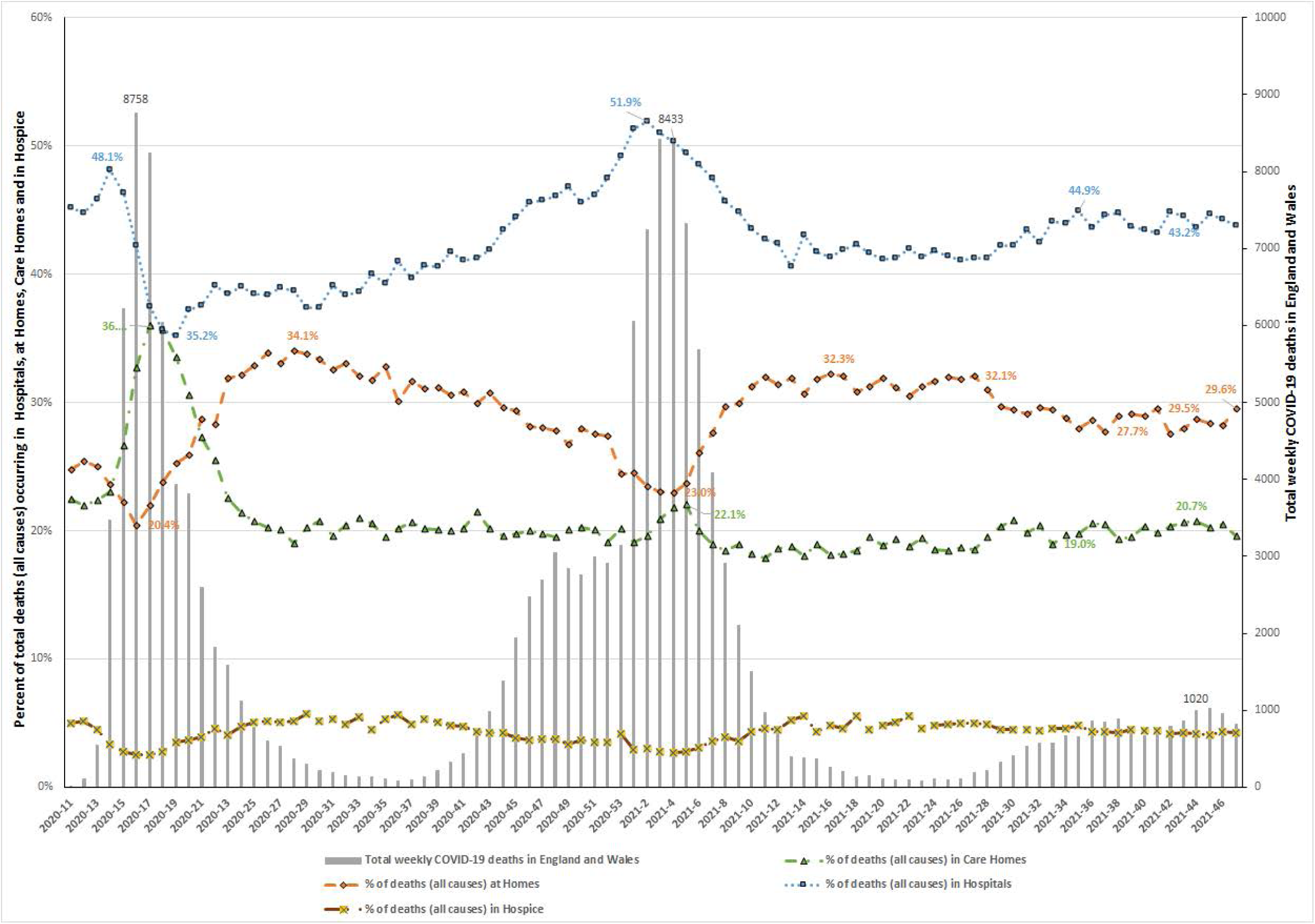
Shows the proportion of weekly deaths in England and Wales (all causes) based on place of death (primary axis) and total weekly COVID-19 deaths (secondary axis). During the 1^st^ wave the Care Homes had worst impact with up to 36% of deaths in England and Wales occurring in Care Homes, associated with decline in deaths occurring the Hospitals. After the sharpest increase in deaths occurring at Care Homes during the 1^st^ waves, the percentage of deaths occurring at Care Homes decreased during the 2^nd^ wave. The deaths occurring at Homes were lowest during the peak weekly deaths of 1^st^ and 2^nd^ waves. However, deaths occurring at home showed sharp increases during the recovery phase of 1^st^ and 2^nd^ waves.

There was also an associated decline in percentage of weekly deaths in hospital from all causes, which subsequently increased during the subsequent surges. The percentage of total deaths occurring at home showed an increase during the recovery phase of the first two surges and remained higher than Care Homes percentage of total deaths during the Delta variant surge.

### The COVID-19 deaths at Care Home; the percentage of Care Home deaths due to COVID-19

Table 3 shows the distribution of the total Care Homes deaths (n=220,092) from all causes and COVID-19 deaths in Care Homes (n=33,329) during the March 7, 2020-November 26, 2021 study period. During the March 7-August 28, 2020 there were 15,414 (20.0%) Care Home deaths due to COVID-19 of the 79,906 total deaths from all causes in care home, and during the August 29, 2020-May 28, 2021 period there was a significant decrease in percentage of Care Home deaths due to COVID-19 comparison with prior period (16,603 COVID-19 deaths [18.5%] of 89,954 Care Home deaths from all causes; RR 0.92; p<0.0001). During the May 29-November 26, 2021 period there was also a significant decrease in Care Home deaths due to COVID-19 compared to prior period (1,312 Care Home deaths due to COVID-19 [2.5%] of the total 53,232 Care Home deaths from all causes; RR 0.13; p<0.0001). Figure 3 illustrate that during the first surge and second surge up to 39.2% and 49% of the total deaths in the Care Homes respectively are due to COVID-19 and during the Delta variant surge, the Care Homes weekly deaths due to COVID-19 are as low as 2.9%, as low as COVID-19 deaths at home. There was also decreased in percentage of deaths due to COVID-19 deaths occurring in Hospitals and Hospice during the Delta variant surge.

**Figure 3:**
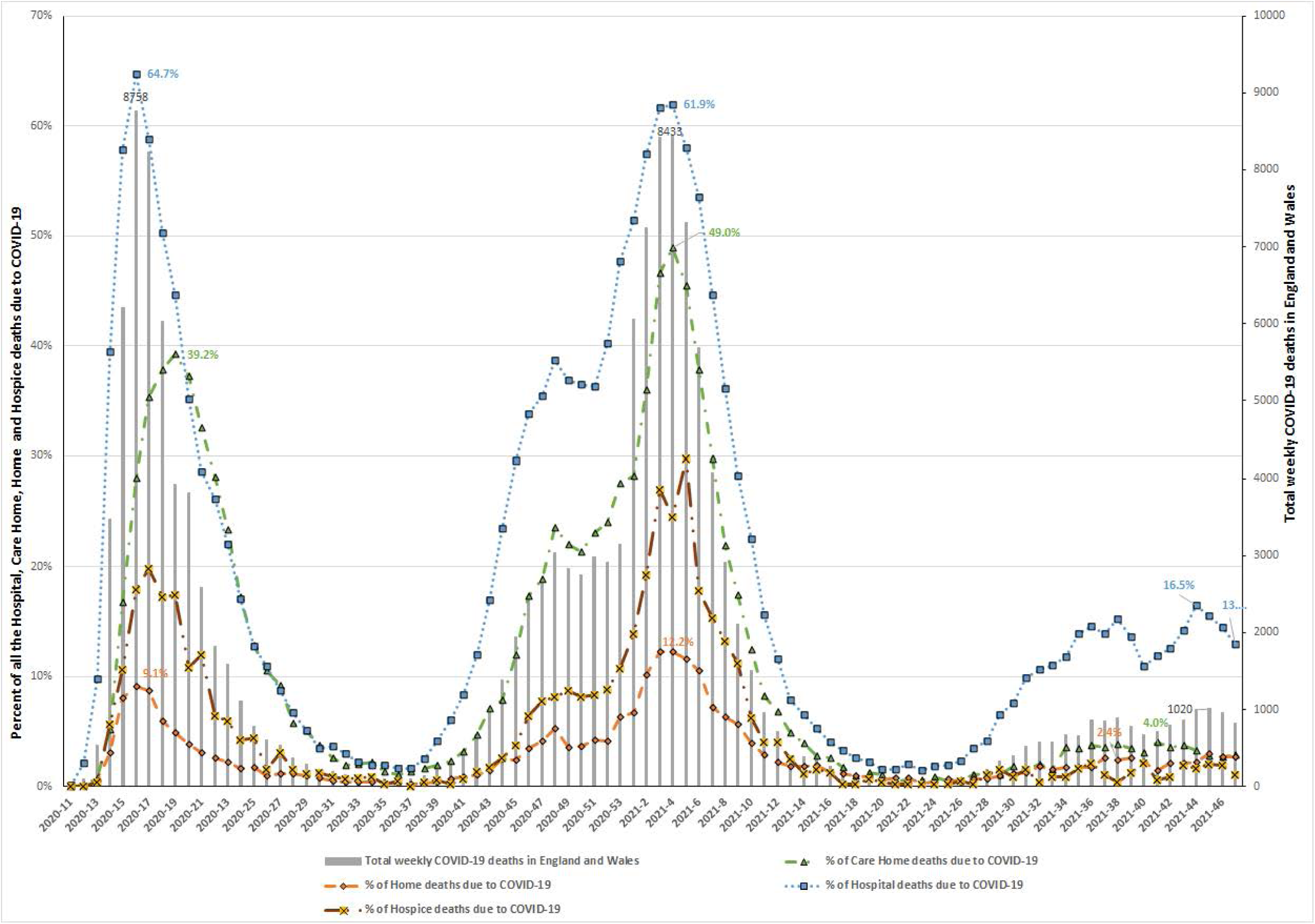
Shows the proportion of weekly deaths due to COVID-19 in England and Wales based on place of death (primary axis) and total weekly COVID-19 deaths (secondary axis). During the 1^st^ and 2^nd^ waves, the COVID-19 deaths were at their highest during the peak weekly deaths in Hospitals, followed by Care Homes, Hospice, and Homes. During the Delta variant surge, the sharpest decline in weekly deaths rates due to COVID-19 was observed in Care Homes, followed by Hospice, Hospitals, and Homes.

### The COVID-19 deaths at Care Home; the impact of Care Home deaths on the total deaths in England and Wales from all causes (Table 4)

**Table 4:**
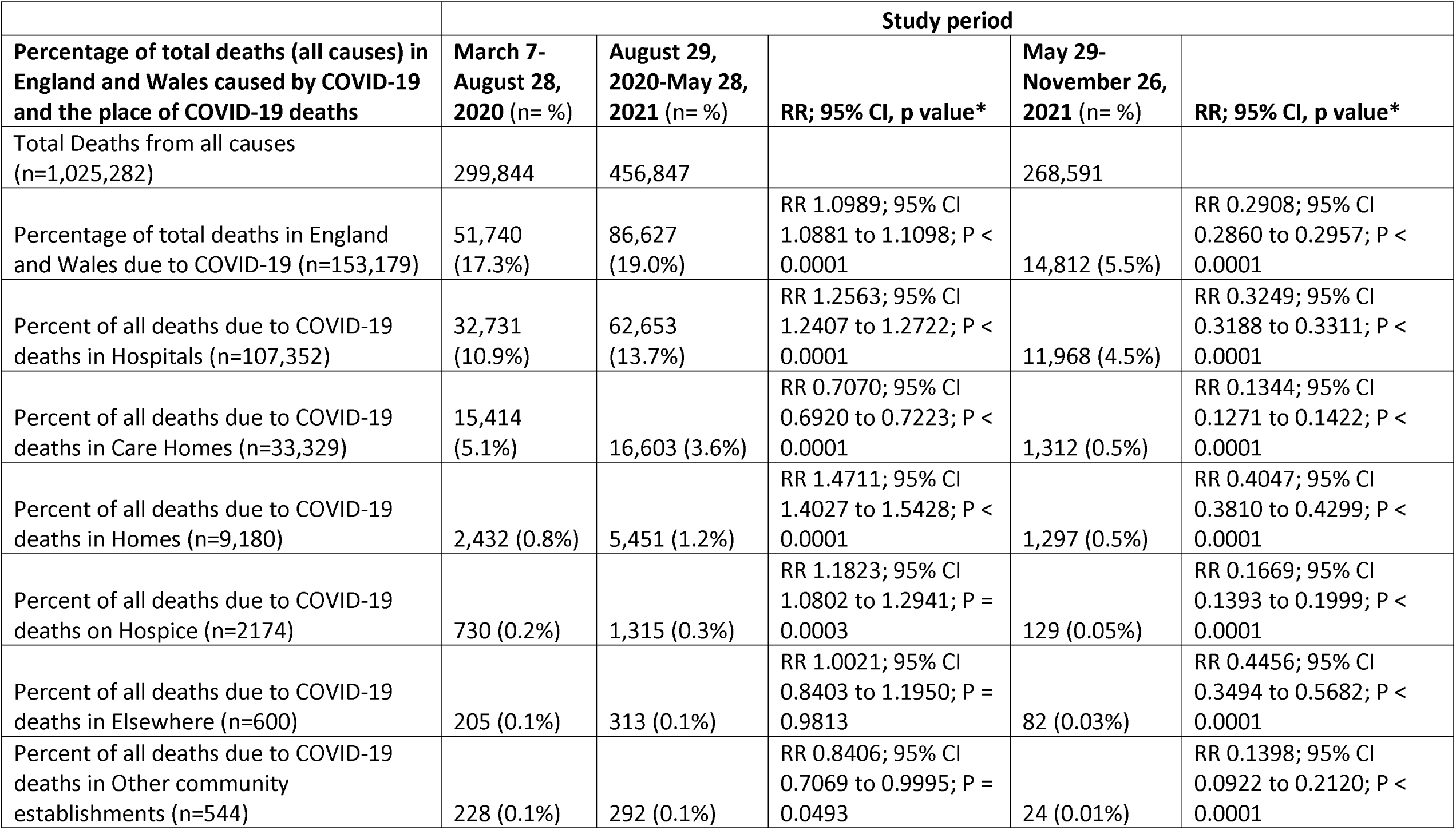
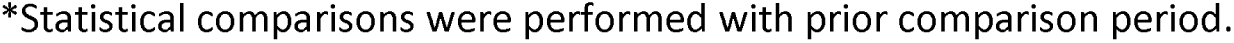
Impact of all COVID-19 deaths, and the COVID-19 deaths at various places on the total deaths (all causes) in England and Wales during the study period (March 7, 2020 -November 26, 2021).

There were 15,414 Care Home COVID-19 deaths (5.1%) of total 299,844 total deaths from all cases in England and Wales during the March 7-August 28, 2020 period and there was a significant decrease in Care Home deaths percentage to 3.6% of the total deaths from all causes (16,603 Care Home COVID-19 deaths/456,847 deaths from all causes; RR 0.70; p<0.0001) during August 29, 2020-May 28, 2021 period. During the Delta variant surge (May 29-November 26, 2021), the percentage of Care Home COVID-19 deaths decreased to 0.5% of total deaths from all causes (1,312 Care Home COVID-19 deaths/268,591 deaths from all causes, RR 0.13; P<0.0001) on comparison with prior period. Figure 4 illustrate the impact of Care Home COVID-19 deaths during the initial surge (March-August 2020), with peak weekly 13.5% of COVID-19 deaths at Care Homes of the total deaths from all causes in England and Wales, during the second surge the percentage of Care Home COVID-19 deaths decreased to 10.7% of the total all cause deaths and during the Delta variant surge, it is at its lowest with Care Home deaths contributing up to 0.8% of total deaths from all cause.

**Figure 4:**
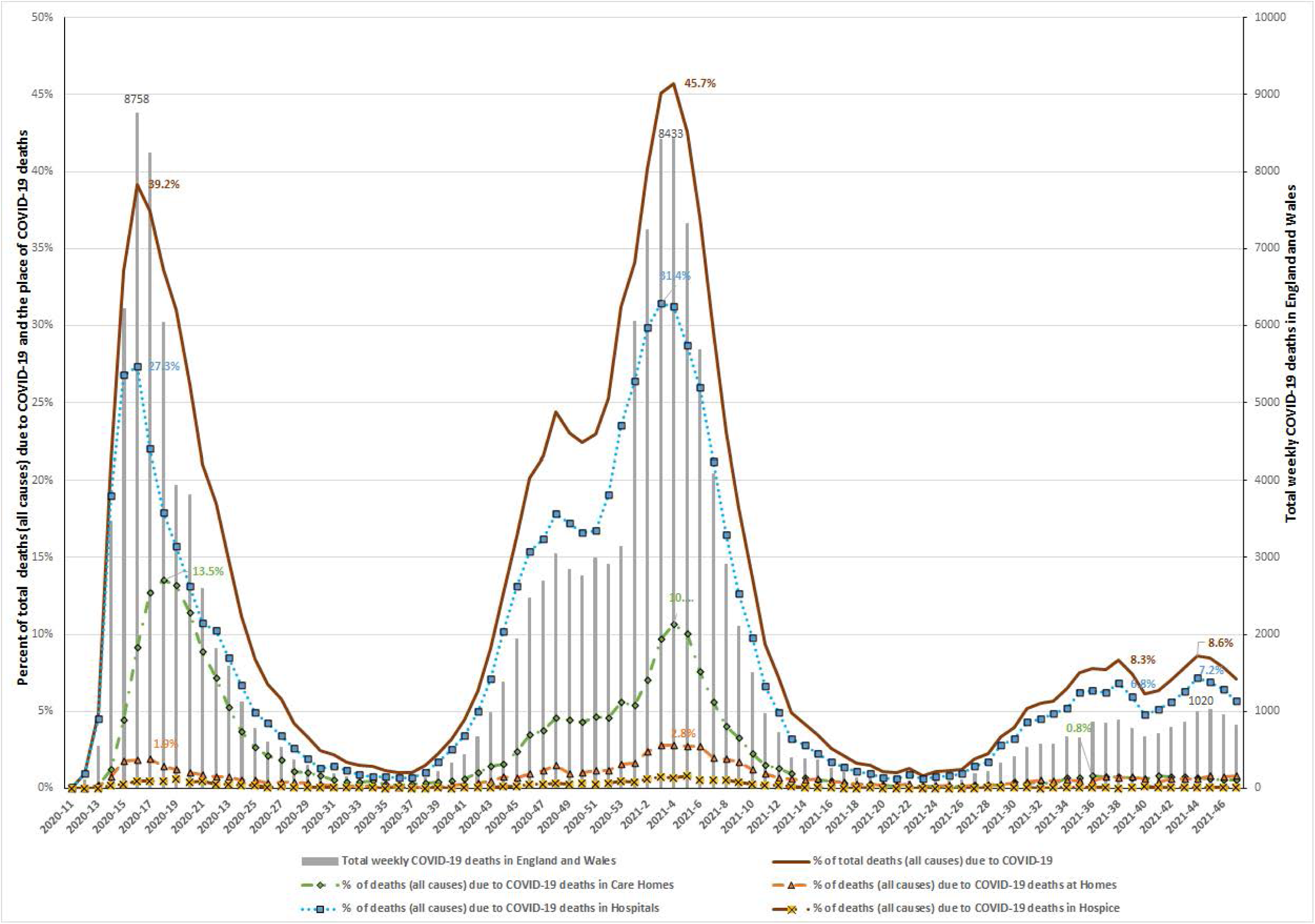
Shows the proportion of weekly total deaths (all causes) due to COVID-19; impact of deaths from COVID-19 at places of death on total deaths (all causes) in England and Wales (primary axis) and total weekly COVID-19 deaths (secondary axis). During the 1^st^ wave up to 39.2% of weekly deaths (all causes) were due to COVID-19 of which up to 27.3% and 13.5% of total deaths are due to COVID-19 in Hospitals and Care Homes respectively. During the 2^nd^ wave up to 45.7% of total deaths (all causes) in England and Wales were due to COVID-19. Hospital weekly COVID-19 deaths increased to as high as 31.4% of deaths from all causes and Care Homes COVID-19 deaths up to 10.7% (declined from 1^st^ wave) of weekly total deaths from all causes. During the Delta variant surge, the weekly COVID-19 deaths declined significantly, with most of the deaths (all causes) due to COVID-19 occurring in the Hospitals.

In summary; during the March 7-August 28, 2020 (first wave), COVID-19 had a major impact on deaths from all causes in England and Wales, with 17.3% of total deaths in England and Wales were due to COVID-19, which increased to 19.0% of the total deaths during the August 29,2020-May 28, 2021 period. During the Delta variant surge, the deaths due to COVID-19 significantly declined with COVID-19 deaths contributing to 5.5% of total deaths from all causes in England and Wales.

The Care Home residents suffered significant mortality during the March 7-August, 28 2020 (first wave) with 29.8% of total COVID-19 deaths occurring in the Care Homes, that resulted in the highest (25.5%) percentage of total deaths from all causes in England and Wales occurring in the Care Homes. During the same period, 20.0% of Care Home deaths occurred due to COVID-19, and the COVID-19 deaths in Care Homes contributed to 5.1% of deaths from all causes in England and Wales. During August 29, 2020-May 28, 2021(the second surge), the percentage of total COVID-19 deaths occurring in the Care Homes significantly decreased (19.2% of the total COVID-19 deaths) with a resultant significant decrease in deaths from all causes occurring in Care Homes (19.7% of total deaths in England and Wales). During the same period, there was a significant decline in the percentage of Care Home deaths that occurred due to COVID-19 (18.5% of all Care Home deaths), and there was also a significant decrease in the percentage of COVID-19 deaths in Care Homes contributing to deaths from all causes in England and Wales (3.6%). During the Delta variant surge (May 29-November 26, 2021) the percent of COVID-19 deaths occurring in Care Homes declined significantly (8.9% of total COVID-19 deaths), the percentage of deaths occurring in Care Homes due to COVID-19 also showed significant decline (2.5% of total deaths in Care Homes) and the Care Homes deaths contributing to total deaths from all causes in England and Wales also significantly decreased (0.5% of total deaths from all causes). These changes in demographic shifts in COVID-19 deaths were associated with a lower percentage of total COVID-19 deaths occurring in the Hospital (63.3%) and the Home (4.7%) during the March-August 2020 first wave. During the subsequent surges (particularly during the Delta variant surge), the percentage of the COVID-19 deaths occurring in the Hospital (80.8%) and Home (8.8%) significantly increased.

## DISCUSSION

The deaths due to COVID-19 declined significantly during the Delta variant surge with COVID-19 deaths contributing to 5.5% of total deaths from all causes (from 17.3% and 19.0% of total deaths from all causes during the 1^st^ and 2^nd^ surges respectively) in England and Wales. This decline in COVID-19 deaths during the Delta variant surge is probably due to less virulence of the Delta variant than prior variants (particularly Alpha variant) as per published data of Public Health England in their regular briefings and potential protective effect of COVID-19 vaccination^17,22–26^.

Our study also showed the dynamic demographic changes in COVID-19 deaths during the SARS-CoV2 pandemic in England and Wales since beginning in March 2021. During the first wave (March 7-August 28, 2020), COVID-19 had a devastating effect on the Care Homes with 29.8% of all COVID-19 deaths occurring in Care Homes, 20% of deaths in Care Home were due to COVID-19 and COVID-19 deaths in Care Homes contributed to 5.1% of deaths from all causes in England and Wales. During the second wave, the total COVID-19 deaths occurring in the Hospitals and Homes significantly increased with a slight, but a significant decrease in COVID-19 deaths occurring in Care Homes.

During the Delta variant surge in our study, the COVID-19 deaths occurring in Care Homes showed a significant sharp decline (from 19.2% to 8.9%) and deaths due to COVID-19 at Care Homes also showed a significant sharp decline (18.5% to 2.5%) since the 2^nd^ surge. There was also a significant increase in COVID-19 deaths occurring at Hospitals (72.3% to 80.8%) and Homes (6.3% to 8.8%) during the Delta variant surge compared to the prior surge.

The reduced deaths rates in the Care Homes during the Delta variant surge are probably due to multiple interventions, first of which is the infection control and protective measures implemented in the Care Homes with the lessons learned from previous surges^9–14^. The greater adoption of COVID-19 vaccination among the Care Home residents and immunity from previous SARS-CoV2 infections are also potential contributary factors for reduced deaths during the Delta variant surge in Care Homes which need further study^17,27–29^.

The data in our study during the first wave is similar to prior reporting of UK Office of National Statistics, and that study highlighted numerical number of deaths in Care Homes during the second wave than the first wave^19^. Our study showed more COVID-19 deaths occurred in all settings including the Care Homes during the second wave, but decreased percentage of COVID-19 deaths occurring in Care Homes with associated decrease in percentages of deaths from all causes occurring in care Homes and deaths due to COVID-19 in Care Homes during the second wave.

The strength of our study is that we performed a comprehensive analysis of nationwide data of deaths from all causes and COVID-19 deaths occurring at various places (Hospitals, Homes, Hospice, other communal establishments and elsewhere) in England and Wales during the SARS-CoV2 pandemic since its beginning in March 2020.

Limitations of our study is that it is an observational study of publicly reported data of the deaths from all causes and COVID-19 in England and Wales during the SARS-CoV2 pandemic and do not involve review of the individual charts to accurately adjudicate the deaths. The other limitations of our study are the generalizability of the findings is limited to the England and Wales population.

Despite the declining COVID-19 deaths observed during the Delta variant surge in our study, the percent of deaths in Care Homes due to COVID-19 during the Delta variant surge is 2.5% which is higher than deaths due COVID-19 at Homes (1.6%). As per the data of 2002, about 0.7% of England and Wales population (n=442,888) living in the Care Homes are at increased risk for mortality as was observed during the first two waves with COVID-19 infections, due to advanced age of the Care Home population (75.6% and 50.3% of Care Home residents are ≥75 years and ≥85 years age groups respectively in England and Wales) and multiple preexisting conditions among the Care Home population^19,30^.

## CONCLUSIONS

In our study, we observed a significantly lower number of COVID-19 deaths occurred in Care Homes and significantly lower number of Care Homes deaths caused by COVID-19 during the Delta variant surge in England and Wales than the prior surges. The Care Home residents are at highest risk for mortality due to advanced age with comorbidities, continuous monitoring and research on COVID-19 preventive interventions is an absolute necessity to further improve the outcomes.

## Data Availability

All data produced in the present work are contained in the manuscript

## Funding source

This research did not receive any specific grant from funding agencies in the public, commercial, or not-for-profit sector

## Ethical Approval statement

None required

## Notes

**Conflicts of interests:** The authors have no conflict of interest or financial relationships relevant to the submitted review to disclose.

**Funding:** None; The author(s) received no financial support for the research, authorship, and/or publication of this article

### Competing Interest Statement

The authors have declared no competing interest.

### Funding Statement

This study did not receive any funding

